# Middle meningeal artery embolization for subdural hematoma: protocol for a systematic review and meta-analysis of randomized controlled trials

**DOI:** 10.1101/2024.02.22.24303232

**Authors:** Alick P Wang, Husain Shakil, Brian J Drake

## Abstract

**Background:** Middle meningeal artery embolization is an emerging neuroendovascular therapy for chronic subdural hematoma. Recently, a number of randomized control trials have been conducted to assess the efficacy of middle meningeal artery embolization to reduce the recurrence or progression of chronic subdural hematoma.

**Methods:** A systematic review will be conducted following the Preferred Reporting Items for Systematic Reviews and Meta-Analysis (PRISMA) guidelines. The authors will systematically search MEDLINE, EMBASE, Cochrane, and ClinicalTrials.gov (National Library of Medicine) for randomized control trials evaluating middle meningeal artery embolization for chronic subdural hematoma. A meta-analysis will be undertaken to compare patients undergoing middle meningeal artery embolization and standard care compared to standard care alone; primary effectiveness endpoints will be symptomatic recurrence, radiographic re-accumulation, or reoperation; secondary safety endpoints will be new disabling stroke, myocardial infarction, or death within 30 days.

**Discussion:** This proposed systematic review and meta-analysis will synthesize and appraise available data regarding middle meningeal artery embolization, a novel neurointerventional therapy. Findings will help clinicians, patients, administrators, policy makers to determine the role of this new treatment and its potential benefits.

**Systematic review registration:** PROSPERO #CRD42024512049

## Background

Chronic subdural hematoma (CSDH) is a common condition, with an incidence of up to 20.6 per 100,000 person-years^1^. It disproportionately affects elderly populations and is often caused by minor trauma. It can cause symptoms ranging from headaches to confusion, decreased level of consciousness, weakness, aphasia, or seizures.

Depending on the volume, local mass effect, and symptoms, chronic SDH has been treated either surgically (burr hole evacuation, craniotomy, or subdural drainage ports) or conservatively (medical therapy or observation/serial imaging). Conservatively managed SDH can progress in up to 34% of cases^2^. After surgical evacuation, recurrence rates have been estimated between 10-20%^3^.

Recently, endovascular embolization of the middle meningeal artery (EMMA) has become an emerging therapy for CSDH^4^. While EMMA was initially used in patients who were at high risk of recurrence, the indications have recently widened to all patients with CSDH. Although EMMA does not immediately remove the SDH or its mass effect, it is hypothesized to devascularize the subdural membrane and prevent recurrence or progression.

EMMA has been used in cases of small, asymptomatic SDH not requiring surgery^5^ as well as in cases of SDH requiring surgical evacuation^6^. A recent systematic review found that EMMA reduced the risk of recurrence from 23.5% to 3.5% (RR 0.17)^7^. However, at the time of publication, there had not been any prospectively conducted randomized control trials (RCTs).

Recently, a number of multi-centre RCTs have been conducted to evaluate the efficacy of EMMA for reducing recurrence or progression in CSDH. A meta-analysis of these trials has yet to be performed.

## Methods/Design

This protocol outlines the design and conduct of our intended systematic review and meta-analysis in compliance with the guideline Preferred Reporting Items for Systematic Reviews and Meta-analyses (PRISMA). The protocol has been submitted to the PROSPERO database and assigned the unique identifier #CRD42024512049. This study was exempt from institutional research ethics board review under article 1B of the University of Toronto policy on Human Research Exempt from REB Review (https://research.utoronto.ca/ethics-human-research/activities-exempt-human-ethics-review)

### Study Aim

We aim to conduct a meta-analysis of available data from RCTs comparing the efficacy of EMMA in addition to standard of care to standard of care alone. The primary efficacy endpoints will be the likelihood of SDH recurrence, re-accumulation, or re-operation at 90 days, and 180 days.

### Data Sources and Study Eligibility

Two authors (APW, HS) will independently search citations from MEDLINE, EMBASE, Cochrane, and ClinicalTrials.gov (National Library of Medicine) without language restriction (see Appendix 1 for search strategy). We will include randomized control trials studying adult patients (≥ 18 years of age) with radiographic diagnosis of chronic or subacute SDH, with at least 10 patients, and at least 90 days of follow-up.

### Treatment Groups

The intervention group of interest will be the use of EMMA in addition to standard of care. The control group will be treatment with standard of care alone. Standard of care will include observation, medical therapy, burr hole, mini-craniotomy, craniotomy, and subdural evacuation port system.

### Outcomes

The primary effectiveness endpoints for this study will be symptomatic recurrence, radiographic re-accumulation, or reoperation at 90, and 180 days. We will also aim to assess a secondary safety endpoint of new major disabling stroke, myocardial infarction, or death from any neurological cause within 30 days.

### Data Extraction

Spreadsheet software will be used by two authors (APW, HS) to extract study design, inclusion/exclusion criteria, randomization characteristics, patient population, primary and secondary outcome measures, and sources of bias, and covariates. Covariates will include age, standard treatment approach used, presence of symptoms, type of embolization material, and use of general anesthetic for EMMA procedure.

### Risk of Bias/Quality Assessment

Two reviewers (APW, HS) will independently perform quality assessment of included studies. The Cochrane ROB 2 tool^8^ will be applied to assess the risk of bias related to the randomization process, deviations from intended interventions, missing data, measurement of the outcome, and selection of the reported results. We judged trials with more than 2 high-risk components as having a moderate overall risk of bias, and trials with more than 4 high-risk components as having a high overall risk of bias. The GRADE approach^9^ will be used for appraisal of the quality of evidence.

### Statistical Analysis

All analyses will be conducted in R version 4.3.1 with a priori specified significance level of p < 0.05 for two-tailed tests. We will follow an intention-to-treat approach in assigning treatment groups.

The odds ratio (OR) of SDH recurrence and associated 95% confidence intervals (CI) for EMMA with standard therapy compared to standard therapy alone will be estimated for each study using logistic regression.

Heterogeneity in the treatment effect across trials will be investigated by the Cochran Q test and measured by the I^2^ statistic, with I^2^ values exceeding 25%, 50%, and 75% representing low, moderate, and high heterogeneity, respectively^10^. Treatment effect estimates will be pooled using a fixed effect Mantel-Haenszel method if heterogeneity is found to be low heterogeneity. A DerSimonian and Laird random-effects model^11^ will be used if moderate-high heterogeneity is found.

Publication bias will be assessed using Funnel plots and Egger’s test^12^. We will further investigate the effect of covariates including age, standard treatment approach, presence of symptoms, history of prior subdural hematoma, use of general anesthetic, and type of embolization material used. We will assess missingness of covariates and outcomes. Based on data availability, we will conduct subgroup analyses based on presence of symptoms, type of standard management (e.g. conservative, medical, burr-hole, and craniotomy), older patients, or patients with prior subdural hematoma. We will not impute data on outcomes or treatment assignment, and will only analyze available data for these variables. For study covariates, we will impute data found to be missing completely at random, or missing at random there is less than 20% missingness.

## Discussion

This proposed systematic review and meta-analysis will address an important knowledge gap regarding EMMA, a novel neurointerventional therapy.

While several studies have evaluated the safety and benefit of EMMA^3^, up until recently there had not been any prospective RCTs conducted to evaluate its efficacy. Recently, a number of multi-centre RCTs have been completed and early results have been presented at the International Stroke Conference 2024.

A meta-analysis of RCTs will help clinicians, patients, administrators, policy makers to determine the role of this new treatment and its potential benefits.

## Data Availability

All data produced in the present study are available upon reasonable request to the authors

## List of abbreviations

CSDH: chronic subdural hematoma
EMMA: embolization of the middle meningeal artery
MMA: middle meningeal artery
PRISMA: Preferred Reporting Items for Systematic Reviews and Meta-analyses
RCT: randomized control trial
SDH: subdural hematoma

## Declarations

### Ethics approval

No ethics approval required for systematic review.

### Availability of data and materials

All data will be extracted from publicly available publications. Extracted data will be made available upon reasonable request.

### Authors’ contributions

APW, HS, and BJD study conception. APW and HS protocol design and drafting.

### Appendix 1

#### Search Strategy

The following search strategy will be employed for the MEDLINE database using the OVID interface.

1. exp Hematoma, Subdural/
2. exp Embolization Therapeutic/
3. exp Meningeal Arteries/
4. exp Endovascular Procedures/
5. Exp Clinical Trial/
6. 2 or 3 or 4
7. 1 and 5 and 6

